# Household factors and the risk of severe COVID-like illness early in the US pandemic

**DOI:** 10.1101/2020.12.03.20243683

**Authors:** Denis Nash, Saba Qasmieh, McKaylee Robertson, Madhura Rane, Rebecca Zimba, Sarah Kulkarni, Amanda Berry, William You, Chloe Mirzayi, Drew Westmoreland, Angela Parcesepe, Levi Waldron, Shivani Kochhar, Andrew R Maroko, Christian Grov, for the CHASING COVID Cohort Study Team

**Affiliations:** Institute for Implementation Science in Population Health (ISPH), City University of New York (CUNY); New York City, New York USA; Department of Environmental, Occupational, and Geospatial Health Sciences, Graduate School of Public Health and Health Policy, City University of New York (CUNY); New York City, New York USA; Department of Maternal and Child Health, Gillings School of Public Health, University of North Carolina, Chapel Hill, NC, USA; Carolina Population Center, University of North Carolina at Chapel Hill, Chapel Hill, NC, USA; Department of Epidemiology and Biostatistics, Graduate School of Public Health and Health Policy, City University of New York (CUNY); New York City, New York USA; Department of Community Health and Social Sciences, Graduate School of Public Health and Health Policy, City University of New York (CUNY); New York City, New York USA

## Abstract

**Objective:** To investigate the role of children in the home and household crowding as risk factors for severe COVID-19 disease.

**Methods:** We used interview data from 6,831 U.S. adults screened for the Communities, Households and SARS/CoV-2 Epidemiology (CHASING) COVID Cohort Study in April 2020.

**Results:** In logistic regression models, the adjusted odds ratio [aOR] of hospitalization due to COVID-19 for having (versus not having) children in the home was 10.5 (95% CI:5.7-19.1) among study participants living in multi-unit dwellings and 2.2 (95% CI:1.2-6.5) among those living in single unit dwellings. Among participants living in multi-unit dwellings, the aOR for COVID-19 hospitalization among participants with more than 4 persons in their household (versus 1 person) was 2.5 (95% CI:1.0-6.1), and 0.8 (95% CI:0.15-4.1) among those living in single unit dwellings.

**Conclusion:** Early in the US SARS-CoV-2 pandemic, certain household exposures likely increased the risk of both SARS-CoV-2 acquisition and the risk of severe COVID-19 disease.

## Introduction

Crowded indoor settings and sustained close contact are associated with an increased likelihood of SARS-CoV-2 spread.^12^ Stay-at-home orders and other non-pharmaceutical measures, such as bans on mass gatherings and physical distancing, were effective in curtailing community transmission.^3,4^ However, these measures may have resulted in shifting the transmission of SARS-CoV-2 to within household settings where high attack rates can occur, with high rates of hospitalization and death.^4–7^ Crowded households can be environments conducive to transmission due to difficulties in maintaining physical distance and effective isolation,^8,9^ and when infected household members have pre-symptomatic or asymptomatic infection.

There is growing evidence suggesting that asymptomatic infections contribute substantially to transmission of SARS-CoV-2 ^10–12^. Younger age may be an important factor driving asymptomatic spread. Studies have shown differences in the presentation of COVID-19 between adults and children, with children less likely than adults to be symptomatic and less likely to present with severe COVID-19 disease.^13–16^ Counterintuitively, other studies have suggested that children may have viral load levels that are comparable to those of adults, and that they could play a role in driving SARS-CoV-2 transmission.^17,18^ One recent study in India found that children and young adults accounted for 30% of cases.^19^ Moreover, the lack of mask use early in the US pandemic indoors among members of the same household who may have been asymptomatic or pre-symptomatic during the first several days of quarantine under stay-at-home orders could have resulted in a higher inoculum and increased disease severity.^20,21,22^

Household studies are important for understanding the role of factors such as household crowding and household age composition on household SARS-CoV-2 transmission. A recent systematic review of 40 SARS-CoV-2 household transmission studies suggests that, while the secondary attack rate within households is high (18.8%, 95% CI 15.4%-22.2%), transmission rates are highest: a) when the primary household cases are symptomatic (19.9%, 95% CI: 14.0% - 25.7%) vs asymptomatic; b) among adult contacts (31%, 95% CI: 19.4% - 42.7%) vs children; and c) in households with only 1 other contact (45.2%, 95% CI 34.1%-51.8) vs those with 3 or more contacts.^2^ Early studies in New York state showed high attack rates, hospitalizations, and deaths within the households of index cases.^6^ And a recent household transmission study conducted in Tennessee and Wisconsin by the CDC found a very high and rapidly occurring secondary infection rate of 53% among household members of an index case, with >70% of secondary cases occurring within 5 days of symptom onset of the index case.^5^ The effect of household transmission versus other community transmission on SARS-CoV-2 severity has not been systematically investigated.

Few household studies have examined the role of children on household transmission of SARS-CoV-2, and those that have relied on small sample sizes.^5,23,24^ Understanding the risk of COVID-19 in crowded households and households with children (regardless of whether they are the primary case in the household) will be important in elucidating the impact of stay-at-home orders and prolonged indoor contact on the risk severe infections. The objective of this study was to examine the effects of household characteristics, primarily the presence of children in the household and household crowding, on the risk of COVID hospitalization during the early phase of the SARS-CoV-2 pandemic in the US.

## Methods

### Study population

Study participants were individuals screened for enrollment into the Communities, Households, and SARS/CoV-2 Epidemiology (CHASING) COVID Cohort study who completed an initial baseline assessment. The CHASING COVID Cohort study is a national prospective cohort study of adults from the US and US territories that was launched on March 28, 2020 to understand the spread and impact of the SARS-CoV-2 pandemic within households and communities. The survey methodology is described in detail in a previous publication.^25^ Briefly, study participants were recruited online through social media platforms or through referrals using advertisements that were in both English and Spanish. The platform Qualtrics (Qualtrics, Provo, UT), an online survey platform widely used in social and behavioral research, was used for data collection.

The initial baseline assessment captured information on household characteristics, underlying risk factors, SARS-CoV-2 symptoms, and health-seeking behaviors such as testing and hospitalizations. A second version of the initial baseline questionnaire was launched on April 9, 2020 to capture healthcare and essential worker status. A total of 6,831 participants had completed an initial cohort screening interview by April 20, 2020. The study protocol was approved by the Institutional Review Board at the City University of New York (CUNY).

### Variable definitions

#### Primary Outcome

The main outcome was self-report of hospitalization for COVID symptoms reported in the two weeks prior to the interview. Symptoms assessed included any of the following: fever, chills, rigors, runny nose, myalgia, headache, sore throat, stomach ache, diarrhea, nasal congestion, nausea, vomiting, cough or coughing up blood or phlegm, shortness of breath. Those reporting any of these symptoms who reported also being hospitalized as a result of their reported symptoms were classified as having the outcome; all other participants were classified as not having the outcome.

#### Primary and secondary exposures

The primary exposure was the presence of any children under 18 years of age living in participants’ household. Secondary exposures were the number of persons living in a household (1, 2-3, more than 4) and the type of property in which the participant lived. Property type was classified as either a multi-unit property (e.g. apartment, condominium, co-op, or building with two or more units), single-unit property (e.g. detached home, or townhouse), or other.

#### Covariates

##### Socio-demographic and behavioral risk factors for COVID

We identified socio-demographic, behavioral and employment factors as confounders of hypothesized exposure-outcome relationships, including age, gender, race/ethnicity, and annual combined income. Additionally, we included potential confounders such as having had close contact with someone who had coronavirus-like symptoms and/or having been involved in the diagnosis or care of someone with confirmed or suspected coronavirus infection. Finally, we considered potential employment-related confounders, including essential worker status, which was defined as having been involved in following roles in the two weeks prior to survey date: healthcare, law enforcement, fire department/first responder, delivery or pick-up services related to food or medications, or in public/private transportation.

#### Community transmission

As community transmission could confound the exposure-outcome relationship, we used lagged population-based, county-level death rates as a proxy for community transmission. We tabulated the number of COVID deaths per 100,000 population for each county using data from the New York Times Github website (from 01/21/2020 to 07/05/2020).^26^ Our proxy for community SARS-CoV-2 transmission was a 5-day moving average of COVID deaths per 100,000 population, lagged by 23 days. Specifically, to use county death rates as a proxy for community transmission in the county, we introduced a lag since COVID deaths follow several other milestones after infection (infection→ incubation→ symptoms→ progression/hospitalization→ death). We assumed that data on the number of deaths for a given day represented community transmission that was occurring 23 days *earlier*, specifically 5 days from infection to symptom onset (reflecting the average incubation period); 5 days from symptom onset to pneumonia; and 13 days from pneumonia diagnosis to death.^27^ For those participants reporting symptoms, we then matched reported timing of symptom onset with community transmission levels 5 days earlier, corresponding to the average incubation period for SARS-CoV-2.^28–30^

#### COVID-related illness

Frequencies of seven measures of COVID-related outcomes were generated to examine the health-seeking behaviors of all participants who 1) reported COVID symptoms; 2) met the CSTE case definition for COVID-like illness which was defined as reporting at least two of following symptoms: fever, chills, myalgia, headache, sore throat, or at least one of the following: cough, shortness of breath^31^; 3) reported seeing or calling a physician or healthcare professional for any of the COVID symptoms they reported, 4) sought but were unable to get a diagnostic test, 5) received diagnostic test, 6) received a laboratory-confirmed diagnosis, or 7) were hospitalized for any of the reported COVID symptoms. All measures were dichotomized as “yes” and “no” with those who reported “do not know” or “not sure” were classified as a “no”.

#### Comorbid conditions

Participants were asked whether they have been told by a health professional that they had heart attack, angina or coronary heart disease, type 2 diabetes, high blood pressure, cancer, asthma, chronic obstructive pulmonary diseases, emphysema, or chronic bronchitis, kidney disease, HIV/AIDS, immunosuppression, and depression.

### Statistical Analysis

Descriptive statistics were generated to examine the socio-demographic, health and behavioral characteristics between households with and without children, and for hospitalized and non-hospitalized participants. Frequencies were generated for all categorical variables and Pearson’s chi-squared test of independence was performed to assess group differences.

A multivariable logistic regression model was used to estimate the association between presence of children in households, number of people living in a household, and property types on the risk of hospitalization with COVID symptoms. We ran three models, all adjusted for age, gender, race/ethnicity, income, close contact, essential worker status, and community transmission rate. Models examining the exposures of the number of people living in household and property types models were also adjusted for presence of children in the household. These variables chosen were based on hypothesized causal associations and confounders, and direct acyclic graphs^32^ were developed for each model.

Given the associations between household crowding with COVID transmission^8,9^, the crude and multivariate associations between 1) presence of children in household or 2) household size on hospitalizations due to COVID were stratified by property type. For each main effect model, we ran a model with an interaction term (i.e., presence of children* property type and household size* property type) and adjusted for the same covariates as the main effects models.

Finally, we described the socio-demographic and behavioral characteristics, as well as comorbidities of participants who were hospitalized with COVID symptoms to those who were not. These included socio-demographic characteristics, and health and behavioral risk factors such as essential worker status, report of having comorbid conditions, and whether participants were in close contact with symptomatic, suspected or confirmed COVID cases. In addition, reported COVID symptoms were examined and ranked for both groups.

### Sensitivity analyses

Three separate sensitivity analyses were performed to assess the potential impact of missing data and misclassification. First, to assess the potential impact of missing values of essential worker status in the initial baseline assessment, a complete case analysis was performed on the participants who had completed the second version of the baseline assessment and for whom essential worker status was known. Given the negative impact of the COVID-19 pandemic on care seeking (including emergency room care)^33^, a second sensitivity analysis assessed the potential impact of excluding persons with COVID symptoms who were not hospitalized in the non-hospitalized group (i.e., differential outcome misclassification). For this analysis we excluded persons who reported symptoms from the denominator of non-hospitalized. The third sensitivity analysis examined the main effects of the exposures on those hospitalized that also had a laboratory confirmed diagnosis for COVID. For this analysis we restricted the outcome to include only those who reported being hospitalized and have received a laboratory-confirmed diagnosis for COVID.

SAS version 9.4 (SAS Institute, Cary, NC) was used for all statistical analyses.

## Results

A total of 6,831 participants completed cohort screening, including 5,348 (78%) who completed the second version of the assessment with the question on essential workers.

### COVID-like illness outcomes

Between March 28, 2020 and April 20, 2020, 58.5% of the study population reported symptoms in the two weeks prior to their study interview, with 25.7% (n=1754) of those meeting the case definition for COVID-like illness^31^ (Figure 1). Twelve percent (n=820) of the study population reported seeing a healthcare provider for these symptoms, 7.6% (n=518) sought SARS-CoV-2 testing but did not receive it, 5.2% (n=357) received a diagnostic test, 2.8% (n=188) received a laboratory confirmed diagnosis of coronavirus, and 2.8% (n=191) reported being hospitalized for COVID-like symptoms (69.6% (n=133) of whom reported laboratory confirmation of their diagnosis, and were considered ‘confirmed’).

**Figure 1:**
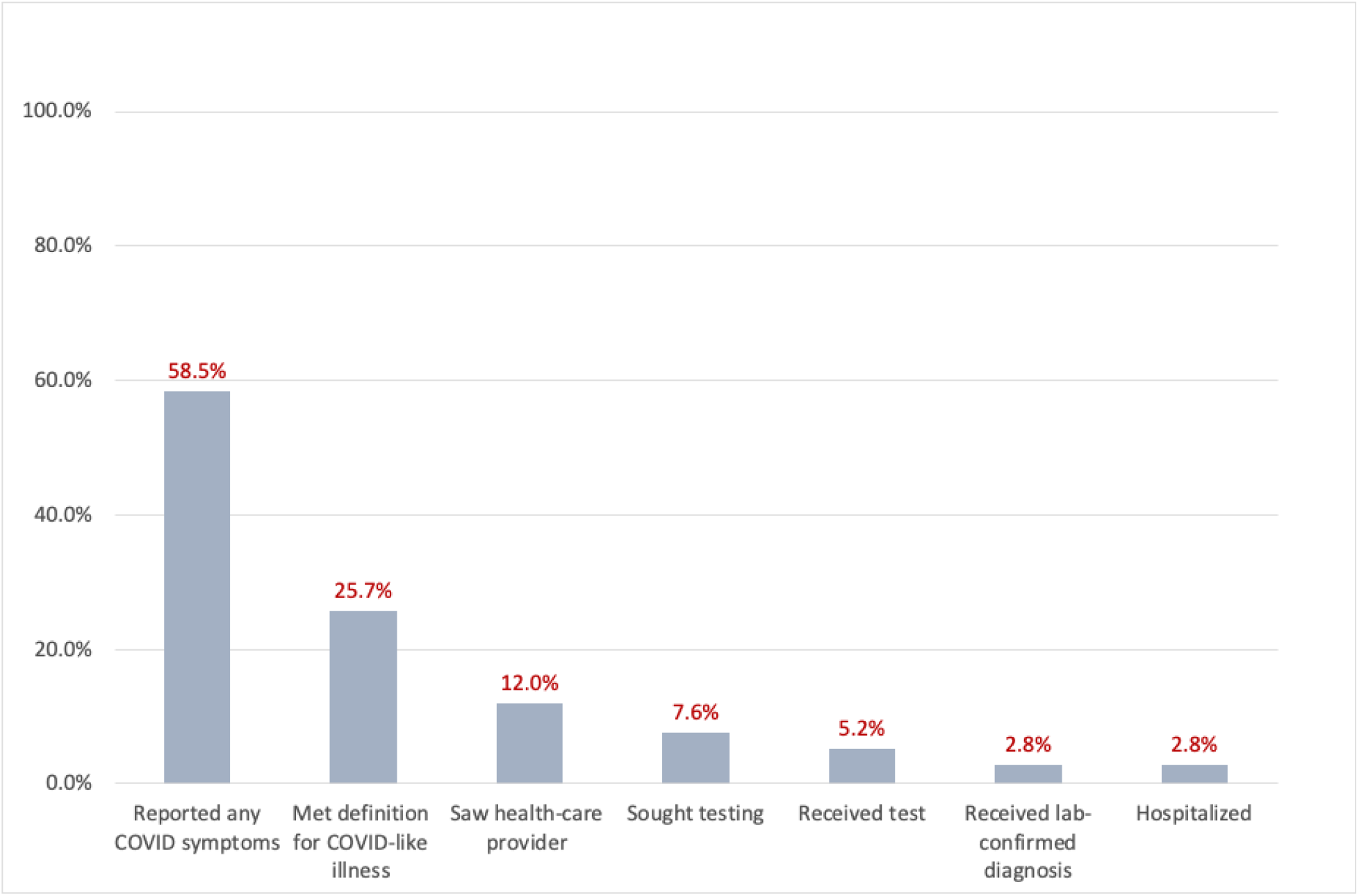
SARS-CoV-2 symptoms among persons screened for enrollment in the CHASING COVID Cohort Study, April 2020

Compared to those without children <18 in the household (Table 1), participants with children were more likely to be under 49 years old (81.2% vs 60.5%), Hispanic (25.9% vs 13.9%), essential workers (26.5% vs 18.8%), and more likely to report having had close contact with someone with coronavirus-like symptoms or a confirmed case (23.3% vs 16.0%). Participants who completed the second version of the assessment were similar to those who completed the first (data not shown).

**Table 1:**
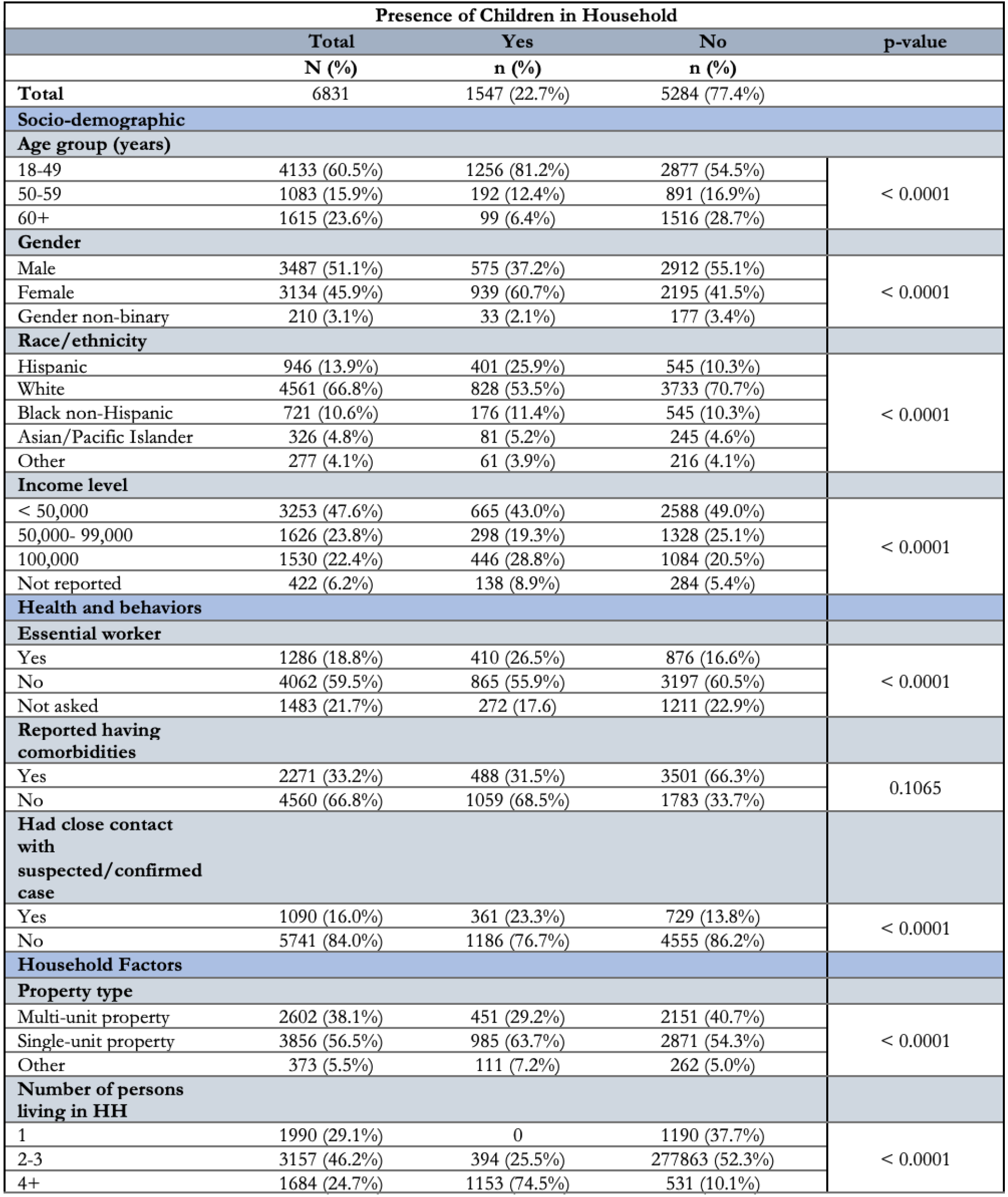
Select socio-demographic, health and behavior characteristics among persons screened for enrollment in the CHASING COVID Cohort (N=6831), April 2020.

| Presence of Children in Household |  |  |  | p-value |
| --- | --- | --- | --- | --- |
|  | Total | Yes | No |  |
|  | N (%) | n (%) | n (%) |  |
| **Total** | 6831 | 1547 (22.7%) | 5284 (77.4%) |  |
| **Socio-demographic** |  |  |  |  |
| **Age group (years)** |  |  |  |  |
| 18-49 | 4133 (60.5%) | 1256 (81.2%) | 2877 (54.5%) | < 0.0001 |
| 50-59 | 1083 (15.9%) | 192 (12.4%) | 891 (16.9%) |  |
| 60+ | 1615 (23.6%) | 99 (6.4%) | 1516 (28.7%) |  |
| **Gender** |  |  |  |  |
| Male | 3487 (51.1%) | 575 (37.2%) | 2912 (55.1%) | < 0.0001 |
| Female | 3134 (45.9%) | 939 (60.7%) | 2195 (41.5%) |  |
| Gender non-binary | 210 (3.1%) | 33 (2.1%) | 177 (3.4%) |  |
| **Race/ethnicity** |  |  |  |  |
| Hispanic | 946 (13.9%) | 401 (25.9%) | 545 (10.3%) | < 0.0001 |
| White | 4561 (66.8%) | 828 (53.5%) | 3733 (70.7%) |  |
| Black non-Hispanic | 721 (10.6%) | 176 (11.4%) | 545 (10.3%) |  |
| Asian/Pacific Islander | 326 (4.8%) | 81 (5.2%) | 245 (4.6%) |  |
| Other | 277 (4.1%) | 61 (3.9%) | 216 (4.1%) |  |
| **Income level** |  |  |  |  |
| < 50,000 | 3253 (47.6%) | 665 (43.0%) | 2588 (49.0%) | < 0.0001 |
| 50,000- 99,000 | 1626 (23.8%) | 298 (19.3%) | 1328 (25.1%) |  |
| 100,000 | 1530 (22.4%) | 446 (28.8%) | 1084 (20.5%) |  |
| Not reported | 422 (6.2%) | 138 (8.9%) | 284 (5.4%) |  |
| **Health and behaviors** |  |  |  |  |
| **Essential worker** |  |  |  |  |
| Yes | 1286 (18.8%) | 410 (26.5%) | 876 (16.6%) | < 0.0001 |
| No | 4062 (59.5%) | 865 (55.9%) | 3197 (60.5%) |  |
| Not asked | 1483 (21.7%) | 272 (17.6) | 1211 (22.9%) |  |
| **Reported having comorbidities** |  |  |  |  |
| Yes | 2271 (33.2%) | 488 (31.5%) | 3501 (66.3%) | 0.1065 |
| No | 4560 (66.8%) | 1059 (68.5%) | 1783 (33.7%) |  |
| **Had close contact with suspected/confirmed case** |  |  |  |  |
| Yes | 1090 (16.0%) | 361 (23.3%) | 729 (13.8%) | < 0.0001 |
| No | 5741 (84.0%) | 1186 (76.7%) | 4555 (86.2%) |  |
| **Household Factors** |  |  |  |  |
| **Property type** |  |  |  |  |
| Multi-unit property | 2602 (38.1%) | 451 (29.2%) | 2151 (40.7%) | < 0.0001 |
| Single-unit property | 3856 (56.5%) | 985 (63.7%) | 2871 (54.3%) |  |
| Other | 373 (5.5%) | 111 (7.2%) | 262 (5.0%) |  |
| **Number of persons living in HH** |  |  |  |  |
| 1 | 1990 (29.1%) | 0 | 1190 (37.7%) | < 0.0001 |
| 2-3 | 3157 (46.2%) | 394 (25.5%) | 277863 (52.3%) |  |
| 4+ | 1684 (24.7%) | 1153 (74.5%) | 531 (10.1%) |  |

### Multivariate analysis

Compared to participants without children in the home, those with children had 4.99 times the adjusted odds (95%CI: 3.16–7.89) in being hospitalized for COVID symptoms (Table 2). No associations were observed between households with more than four persons and hospitalization for COVID symptoms (aOR:1.11; 95%CI: 0.49–2.47) compared to one-person households. Participants who lived in a multi-unit property compared to those living in a single-unit had 4.62 (95%CI: 2.79–7.66) times higher adjusted odds of being hospitalized.

**Table 2:**
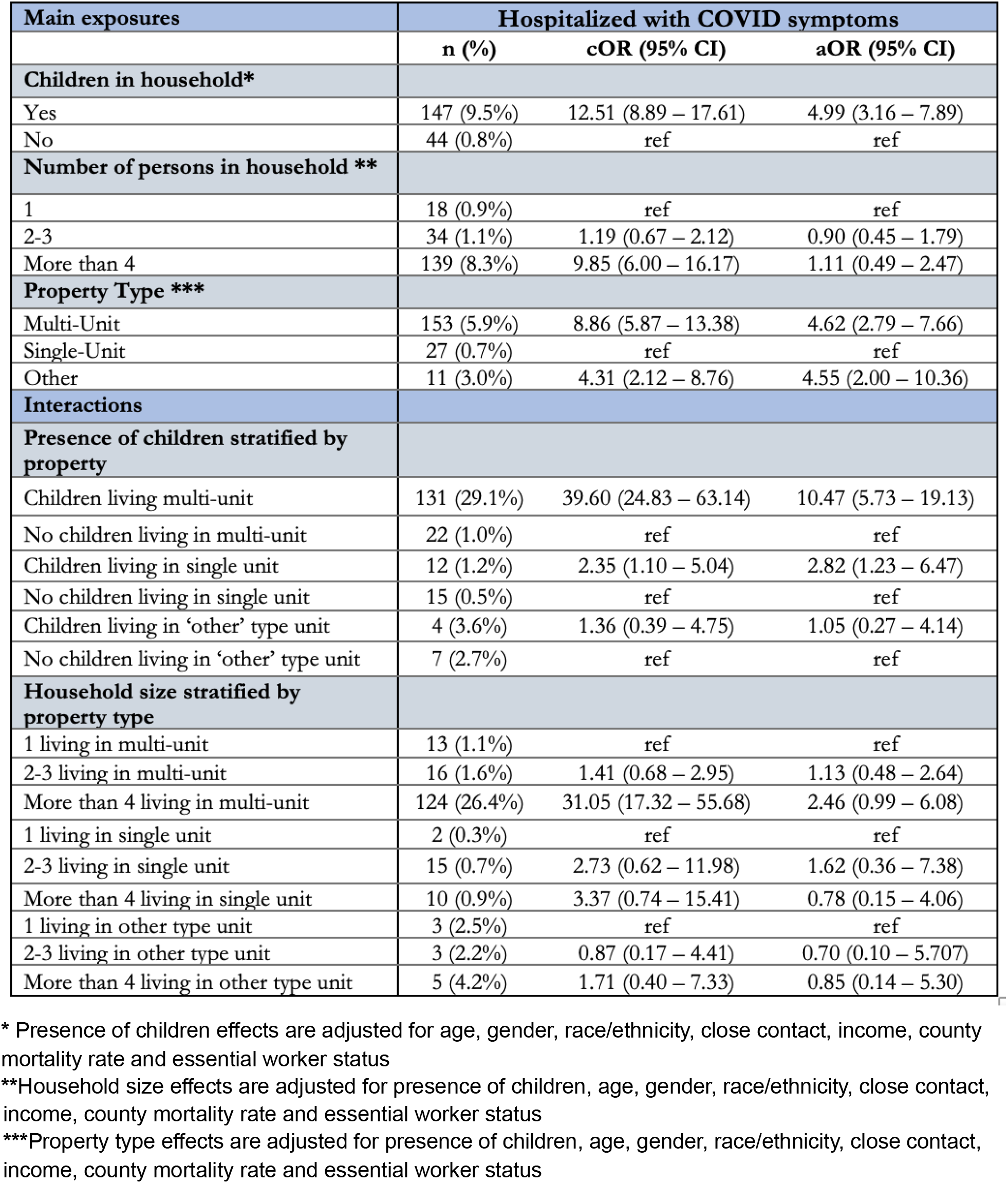
Main effects of presence of children, number of persons in households and property type on hospitalization for COVID-like symptoms (N=6831), April 2020.

| Main exposures | Hospitalized with COVID symptoms |  |  |
| --- | --- | --- | --- |
|  | n (%) | cOR (95% CI) | aOR (95% CI) |
| **Children in household\*** |  |  |  |
| Yes | 147 (9.5%) | 12.51 (8.89 – 17.61) | 4.99 (3.16 – 7.89) |
| No | 44 (0.8%) | ref | ref |
| **Number of persons in household \*\*** |  |  |  |
| 1 | 18 (0.9%) | ref | ref |
| 2-3 | 34 (1.1%) | 1.19 (0.67 – 2.12) | 0.90 (0.45 – 1.79) |
| More than 4 | 139 (8.3%) | 9.85 (6.00 – 16.17) | 1.11 (0.49 – 2.47) |
| **Property Type \*\*\*** |  |  |  |
| Multi-Unit | 153 (5.9%) | 8.86 (5.87 – 13.38) | 4.62 (2.79 – 7.66) |
| Single-Unit | 27 (0.7%) | ref | ref |
| Other | 11 (3.0%) | 4.31 (2.12 – 8.76) | 4.55 (2.00 – 10.36) |
| **Interactions** |  |  |  |
| **Presence of children stratified by property** |  |  |  |
| Children living multi-unit | 131 (29.1%) | 39.60 (24.83 – 63.14) | 10.47 (5.73 – 19.13) |
| No children living in multi-unit | 22 (1.0%) | ref | ref |
| Children living in single unit | 12 (1.2%) | 2.35 (1.10 – 5.04) | 2.82 (1.23 – 6.47) |
| No children living in single unit | 15 (0.5%) | ref | ref |
| Children living in 'other' type unit | 4 (3.6%) | 1.36 (0.39 – 4.75) | 1.05 (0.27 – 4.14) |
| No children living in 'other' type unit | 7 (2.7%) | ref | ref |
| **Household size stratified by property type** |  |  |  |
| 1 living in multi-unit | 13 (1.1%) | ref | ref |
| 2-3 living in multi-unit | 16 (1.6%) | 1.41 (0.68 – 2.95) | 1.13 (0.48 – 2.64) |
| More than 4 living in multi-unit | 124 (26.4%) | 31.05 (17.32 – 55.68) | 2.46 (0.99 – 6.08) |
| 1 living in single unit | 2 (0.3%) | ref | ref |
| 2-3 living in single unit | 15 (0.7%) | 2.73 (0.62 – 11.98) | 1.62 (0.36 – 7.38) |
| More than 4 living in single unit | 10 (0.9%) | 3.37 (0.74 – 15.41) | 0.78 (0.15 – 4.06) |
| 1 living in other type unit | 3 (2.5%) | ref | ref |
| 2-3 living in other type unit | 3 (2.2%) | 0.87 (0.17 – 4.41) | 0.70 (0.10 – 5.707) |
| More than 4 living in other type unit | 5 (4.2%) | 1.71 (0.40 – 7.33) | 0.85 (0.14 – 5.30) |

\* Presence of children effects are adjusted for age, gender, race/ethnicity, close contact, income, county mortality rate and essential worker status
\*\*Household size effects are adjusted for presence of children, age, gender, race/ethnicity, close contact,
income, county mortality rate and essential worker status \*\*\*Property type effects are adjusted for presence of children, age, gender, race/ethnicity, close contact, income, county mortality rate and essential worker status

When stratified by property type, compared to participants without children living in a multi-unit property, participants with children living in multi-unit property had 10.47 times the adjusted odds for being hospitalized for those symptoms (95%CI: 5.73–19.13). Participants living in households with more than 4 people in multi-unit property had 2.46 times the adjusted odds (95%CI: 0.99–6.08) of being hospitalized with COVID symptoms than participants living alone in a multi-unit property. No other statistically significant associations were observed between household sizes on hospitalization when stratified by property type.

### Characteristics of hospitalized participants compared to those not hospitalized

A total of 191 participants were hospitalized for their reported symptoms (Table 3). Compared with all other participants (n=6,638), those hospitalized were more likely to be between the age of 18 and 49 years of age (84.4% vs 59.8%), male (82.2% vs 50.2%), and Hispanic (70.7% vs 12.2%). Hospitalized participants were more likely to report a comorbid condition (85.4% vs 31.7%) and to have had contact with a symptomatic case or suspected or confirmed case (76.4% vs 14.2%). They were also more likely to be essential workers (80.1% vs 17.1%). Fever and sore throat were the two most commonly reported symptoms which were reported by 71.7% and 70.2% of participants, respectively. About 76.4% of hospitalized participants received a test, and 69.6% of them received a laboratory-confirmed diagnosis.

**Table 3:** Characteristics of participants by status of hospitalization with COVID-like symptoms, April 2020.

|  | Hospitalized with COVID-symptoms |  |  |
| --- | --- | --- | --- |
| Socio-demographic Factors | Yes<br>n (%) | No<br>n (%) | p-value |
| **Age group (years)** |  |  |  |
| 18-49 | 162 (84.8%) | 3969 (59.8%) | <0.0001 |
| 50-59 | 8 (4.2%) | 1075 (16.2%) |  |
| 60+ | 21 (11.0%) | 1594 (24.0%) |  |
| **Gender** |  |  |  |
| Male | 157 (82.2%) | 3329 (50.2%) | <0.0001 |
| Female | 31 (16.2%) | 3102 (46.7%) |  |
| Non-binary | 3 (1.6%) | 207 (3.1%) |  |
| **Race/ethnicity** |  |  |  |
| Hispanic | 135 (70.7%) | 811 (12.2%) | <0.0001 |
| White | 29 (15.2%) | 4531 (68.3%) |  |
| Black non-Hispanic | 19 (10.3%) | 702 (10.6%) |  |
| Asian/Pacific Islander | 2 (1.1%) | 324 (4.9%) |  |
| Other | 6 (3.1%) | 270 (4.1%) |  |
| **Income level** |  |  |  |
| < 50,000 | 50 (26.2%) | 3202 (48.2%) | <0.0001 |
| 50,000- 99,000 | 14 (7.3%) | 1612 (24.3%) |  |
| 100,000 | 123 (64.4%) | 1407 (21.2%) |  |
| Missing | 4 (2.1%) | 417 (6.3%) |  |
| **Health and behavioral Factors** |  |  |  |
| **Reported having comorbidities** |  |  |  |
| Yes | 165 (85.4%) | 2105 (31.7%) | <0.0001 |
| No | 26 (13.6%) | 4533 (68.3%) |  |
| **Had contact with symptomatic or suspected/confirmed case** |  |  |  |
| Yes | 146 (76.4%) | 944 (14.2%) | <0.0001 |
| No | 45 (23.6%) | 5694 (85.8%) |  |
| **Essential Worker** |  |  |  |
| Yes | 153 (80.1%) | 1133 (17.1%) | <0.0001 |
| No | 29 (15.2%) | 4032 (60.7%) |  |
| Not asked | 9 (4.7%) | 1473 (22.2%) |  |

| **Reported symptoms (ranked)** |  |  |  |
| --- | --- | --- | --- |
| Fever | 137 (71.7%) | 314 (4.7%) | <0.0001 |
| Sore throat | 134 (70.2%) | 924 (13.9%) | <0.0001 |
| Headache | 32 (16.8%) | 1867 (28.1%) | 0.0005 |
| Myalgia | 31 (16.2%) | 758 (11.4%) | 0.040 |
| Cough with phlegm | 30 (15.7%) | 831 (12.5%) | 0.191 |
| Shortness of breath | 28 (14.7%) | 571 (8.6%) | 0.004 |
| New cough | 27 (14.4%) | 769 (11.6%) | 0.279 |
| Runny nose | 26 (13.6%) | 1641 (24.7%) | 0.0004 |
| Diarrhea | 22 (11.5%) | 853 (12.9%) | 0.587 |
| Nasal congestion | 21 (11.0%) | 1415 (21.3%) | 0.0006 |
| Chills | 18 (9.4%) | 360 (5.4%) | 0.017 |
| Nausea | 15 (7.9%) | 402 (6.1%) | 0.306 |
| Vomit | 9 (4.7%) | 82 (1.2%) | <0.0001 |
| Cough with blood | 3 (1.6%) | 19 (0.3%) | 0.002 |
| **Saw healthcare provider for symptoms** |  |  |  |
| Yes | 173 (90.6%) | 646 (9.7%) | <0.0001 |
| No | 18 (9.4%) | 5992 (90.3%) |  |
| **Testing status** |  |  |  |
| Sought test | 18 (9.4%) | 500 (7.5%) | <0.0001 |
| Received test | 146 (76.4%) | 211 (3.2%) |  |
| Did not need or try | 27 (14.1%) | 5927 (89.3) |  |
| **Received lab-confirmed diagnosis** |  |  |  |
| Yes | 133 (69.6%) | 55 (0.8%) | <0.0001 |
| No | 58 (30.4%) | 6583 (99.2%) |  |

## Discussion

Our study suggests that certain household exposures at the beginning of the SARS-CoV-2 pandemic in the US not only increased the risk of SARS-CoV-2 acquisition, but also increased the risk of severe COVID-19 disease, requiring hospitalization. Household crowding and having children in the home were both strong and independent risk factors for being hospitalized with SARS-CoV-2 early in the US pandemic. Our findings have implications for public health recommendations in areas and during times where the risk of household transmission of SARS-CoV-2 may be higher, such as immediately prior to and after issuing stay at home orders, when the point prevalence of SARS-CoV-2 in the affected communities may be at its highest level. Essential workers, families with children, and those living in crowded indoor settings may be at particularly high risk for being hospitalized with SARS-CoV-2, and tailored recommendations to reduce the risk of household transmission are needed when community transmission is high.

Household transmission occurs rapidly after an index case introduces the infection and with high household attack rates, can originate from both children and adults^19^, and are associated with high rates of hospitalization and death.^2,5,6^

Early in the US pandemic, even when mask use was recommended and became the norm outside the home, and even in areas where stay at home orders had been put in place, mask use inside the home or other at-home risk mitigation measures were not recommended except in situations where there were ill/infected persons (when 10-14 days of isolation using a separate bedroom and bathroom was recommended to reduce household transmission). However, because of household crowding, a lack of space in the households, or a need to go to work, provide childcare, or care for other household/family measures, isolation of ill/infected persons and quarantine of those who have had a high risk exposure (e.g., to a confirmed case) is not always feasible, and other risk mitigation measures (e.g., mask wearing, opening windows) are therefore needed.

A higher infectious dose can result in a more severe course of SARS-CoV-2 infection. Mask use is an effective strategy to both reduce the risk of onward spread from an infected person to susceptibles, and also reduces the risk of infection to the mask wearer.^34^ However, infections still can occur when masks are being used by infected and susceptible persons, but these infections may be more likely to result in asymptomatic or milder SARS-CoV-2 infection, because of a lower infectious dose.^20^ It is therefore possible that a lack of mask use at home during quarantine and isolation results in a higher infectious dose of SARS-CoV-2 than would occur with masks, especially in smaller homes, with more household members, including children.

Stay at home orders usually go into effect when community transmission (and point prevalence) is highest, and by definition, they increase the amount of time that household members spend together indoors. Given this, the high attack rates of SARS-CoV-2 within households, and the higher severity of SARS-CoV-2 infections that can result from prolonged unmasked, indoor exposures when entire households are quarantined, public health officials should consider recommending mask use and other risk mitigation strategies (e.g., open windows, reduced close contact, frequent at-home testing) in all households with more than one person for a period of time immediately before and after stay at home orders go into effect.

Essential workers have several potential risks for SARS-CoV-2 infection, including commuting and exposure while at work, but they also may be at higher risk for acquiring or transmitting infection at home as well. In NYC and Chicago, a COVID-19 hotspot analysis found that hot spots had more household crowding, in NYC, and tended to be middle income, working class neighborhoods that may have higher concentrations of essential workers.^9^

Our study, which included substantial numbers of essential workers who were hospitalized, looked within households and described some of the risk factors for severe COVID-19 early in the US pandemic. Essential workers with young children need childcare, which could put their children at risk of infection. Children with SARS-CoV-2 infection have high enough viral load to transmit to others, and may be more likely to have unrecognized infections. When children become ill, isolation may be even more challenging than for adults, and adults providing care for them are at higher risk, possibly of a more severe SARS-CoV-2 infection, especially when it is difficult to follow infection control practices, such as mask wearing at home by all household members.

Our study has limitations worth noting. First, this was not a household transmission study, and therefore we could not pinpoint household transmission as the likely source of the infection that resulted in infection and hospitalization among our study participants. We therefore cannot say whether and the extent to which household transmission occurred, and if it did, when a child in the household was the source of infection to our study participants. We also did not assess participants’ mask use at home. Finally, while we controlled for several possible confounders of the association of household crowding and children in the home with SARS-CoV-2 risk, unmeasured confounding could partially explain our observed associations.

Our study also had some strengths. As a large epidemiologic cohort study, it was possible to examine the association of several potentially important household-level risk factors with a relatively rare outcome of SARS-CoV-2 hospitalization, while controlling for potential confounding factors. We also had a geographically diverse sample that included several essential workers. Finally, our findings were robust to the three sensitivity analyses, which generated similar findings to the main analysis.

## Conclusions

Early in the US SARS-CoV-2 pandemic, certain household exposures likely increased the risk of both SARS-CoV-2 acquisition and the risk of severe COVID-19 disease requiring hospitalization. These findings may have implications for mask wearing and other mitigation strategies at home immediately prior to and immediately after ‘stay at home’ orders go into effect.

## Data Availability

Please contact study team.

https://cunyisph.org/chasing-covid/

## Acknowledgements

We would like to acknowledge the CHASING COVID Cohort Study participants for their contributions to this research.

